# HEARING MORE TO HEAR LESS: A SCOPING REVIEW OF HEARING AIDS FOR TINNITUS RELIEF

**DOI:** 10.1101/2021.03.08.21253134

**Authors:** Laure Jacquemin, Annick Gilles, Giriraj Singh Shekhawat

**Affiliations:** University Department of Otorhinolaryngology and Head & Neck surgery, Antwerp University Hospital, Edegem, Belgium; Department of Translational Neurosciences, Faculty of Medicine and Health Sciences, Antwerp University, Wilrijk, Belgium; Department of Education, Health & Social Work, University College Ghent, Ghent, Belgium; College of Nursing and Health Sciences, Flinders University, Bedford Park, SA, Australia; Tinnitus Research Initiative, Regensburg, Germany; Ear Institute, University College London, UK

**Keywords:** hearing aids, sound amplification, tinnitus, scoping review, treatment

## Abstract

**Background:** Tinnitus, the perception of a sound in absence of an external auditory source, can significantly impact ones’ quality of life. As tinnitus is often associated with hearing loss, hearing aids have been proposed for tinnitus relief in literature for more than 70 years. While there is a long history of clinical work and research on this topic, there is a need for recent literature to be reviewed and guide decision making in tinnitus management.

**Objective:** The aim of this scoping review is to provide an update of the available evidence on hearing aids for tinnitus, focusing on the effect of sound amplification, to draw conclusions for clinical practice and identify gaps in the field. A consultation exercise was included to discuss current issues that practitioners and carers themselves face but remain under-researched.

**Design:** This scoping review was conducted based on the six-stage framework of Arksey et al. (2005). Studies were included if they investigated hearing aids for tinnitus and were published after 2011. Databases of PubMed and Scopus were explored on the 16th of November 2020. The search was limited to English manuscripts. A total of 28 primary research studies were selected.

**Results:** Positive results of hearing aids for tinnitus relief were shown by 68 % of the studies, whereas 14 % demonstrated no change in tinnitus perception. As the quality of the evidence across studies was variable, no consensus can be reached regarding the use of hearing aids as a treatment for tinnitus. Nevertheless, recent studies were more likely to focus on optimizing the effect of hearing aids and better predicting which tinnitus patients benefit from hearing aids. The experts stated that the findings were in agreement with their view on the scientific evidence and they emphasized the importance of reaching consensus.

**Conclusions:** The majority of the studies supported the use of hearing aids for tinnitus relief. Hence, there was some scientific support for it, but the quality of evidence was questioned. Stronger methodology in future studies is needed to reach consensus and support clinical guidelines development.

## Introduction

Tinnitus is the perception of a sound (e.g. sizzling, ringing or hissing) in the absence of a corresponding acoustic source (Baguley et al. 2013; Jastreboff 1990). It is a common experience (10-15% in adults) which, for some (2.4% in adults), may cause a considerable amount of distress and decreased quality of life (Axelsson et al. 1989; Baguley et al. 2013). As tinnitus has no cure and is generally associated with hearing loss, hearing aids have been proposed for tinnitus relief for more than 70 years (Saltzman et al. 1947). In 2013, Shekhawat et al. conducted a scoping review to explore the role of hearing aids in tinnitus management (Shekhawat, Searchfield and Stinear 2013). The authors concluded that some evidence had been provided for the use of hearing aids. However, the authors identified a need for stronger methodology and randomized control trials.

The exact working mechanism behind tinnitus relief with hearing aids is not yet fully understood. Proposed mediation factors include reduction in central gain, habituation, reduction in communication stress, altered neuronal plasticity and masking (Beck 2011; Robert W. Sweetow et al. 2010). The maladaptive neuronal gain in the central auditory system as a response to the cochlear deafferentation (i.e. central gain) might be reduced as hearing aids provide additional sensory input which was lost (Noreña 2011). This repaired input can also lead to increased neuronal activity in the auditory pathway, possibly interfering with the central processing of tinnitus (Jastreboff et al. 1993). As such, the contrast between tinnitus and background stimuli can be reduced, facilitating habituation to tinnitus. Decreased communication stress, on the other hand, as a result of hearing aids use, can support coping with tinnitus (Surr et al. 1985). A long term effect of hearing aids that has been proposed through animal studies is reversing the tinnitus-related cortical reorganization (Eggermont 2008). Finally, the model of masking suggests that hearing aids can (partially) mask the tinnitus percept and, thus, divert attention away from it to more meaningful sounds (Coles et al. 1987).

Masking can also be achieved by providing additional, more acceptable stimuli with a sound generator. This concept was introduced in 1976 by Vernon et al. (J. Vernon et al. 1976). Beyond masking, the potential positive effect of sound generators for tinnitus might be due to a sense of relief, habituation, neuromodulation or relaxation (Jastreboff and Hazell 1993; Noreña et al. 2005; R. W. Sweetow et al. 2010; Tass et al. 2012; J. A. Vernon et al. 2003). However, as sound therapy is often part of a more extensive tinnitus management program, such as Tinnitus Retraining Therapy (Jastreboff et al. 2000) or bimodal therapy (Luyten & Jacquemin et al. 2020), it has been difficult to draw conclusions concerning its isolated effectiveness (Sereda et al. 2018). The two approaches of amplification and sound generation can be incorporated in combination devices.

Previous reviews investigating the effect of sound-enriching devices (i.e. hearing aids, sound generators, combination devices) reached similar conclusions: positive results have been demonstrated in terms of tinnitus relief, however the quality of evidence is low (Hoare et al. 2014; Hobson et al. 2012; Sereda et al. 2018; Tutaj et al. 2018). Hence, it seems that from 2012 until 2018, no considerable improvement has been made in this field, and as such, conclusive evidence is absent.

The current scoping review provides an overview of all available evidence on hearing aids for tinnitus published after the scoping review by Shekhawat et al. in 2013, focusing mainly on the effect of sound amplification itself. Over the last years, considerable amount of research has been conducted in this field. As such, this scoping review might lead to new insights. A last, but essential, stage of a scoping review will be included (and was missing in Shekhawat’s review), namely a consultation exercise which discusses current issues that practitioners and carers themselves face but remain under-researched. Finally, this type of review will make it possible to draw conclusions for clinical practice and identify gaps in the field from the data of different types of studies (e.g. randomized controlled trials (RCTs), retrospective studies, clinical gathered data), thus without excluding studies based on their quality (Arksey and O’Malley 2005; Mays et al. 2001).

## Methods

A scoping review was conducted according to the framework of Arksey and O’Malley (2005). In order to collect all relevant primary research studies published on this topic since 2011, an advanced search was carried out using the databases of PubMed and Scopus on the 16 ^th^ of November 2020, limited to English manuscripts. The title and/or abstract had to include the key words ‘tinnitus’ and ‘hearing aid’. In figure 1, the details of the search strategy are presented.

**Figure 1:**
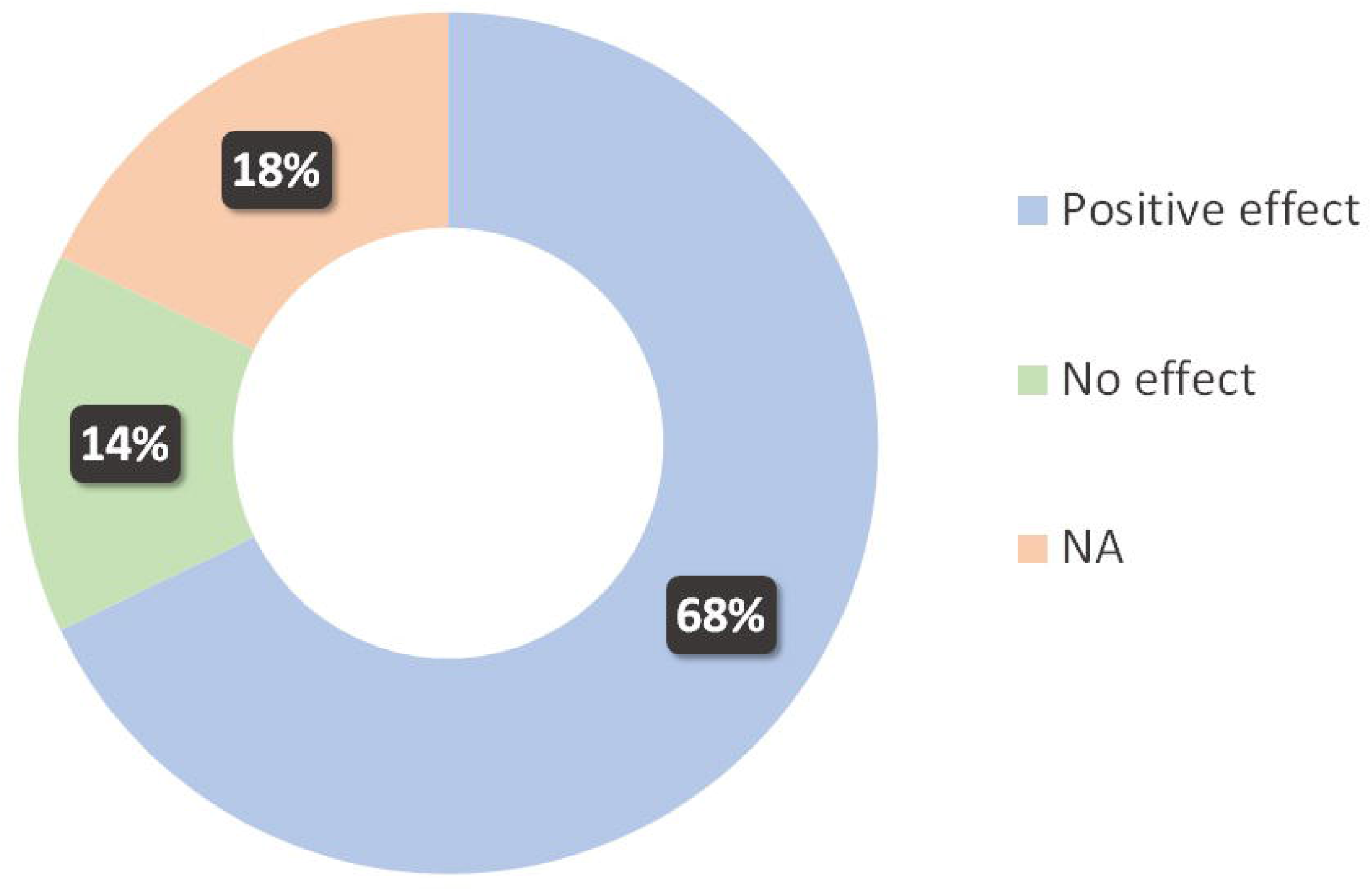
Flowchart of the study selection

**Figure 2:**
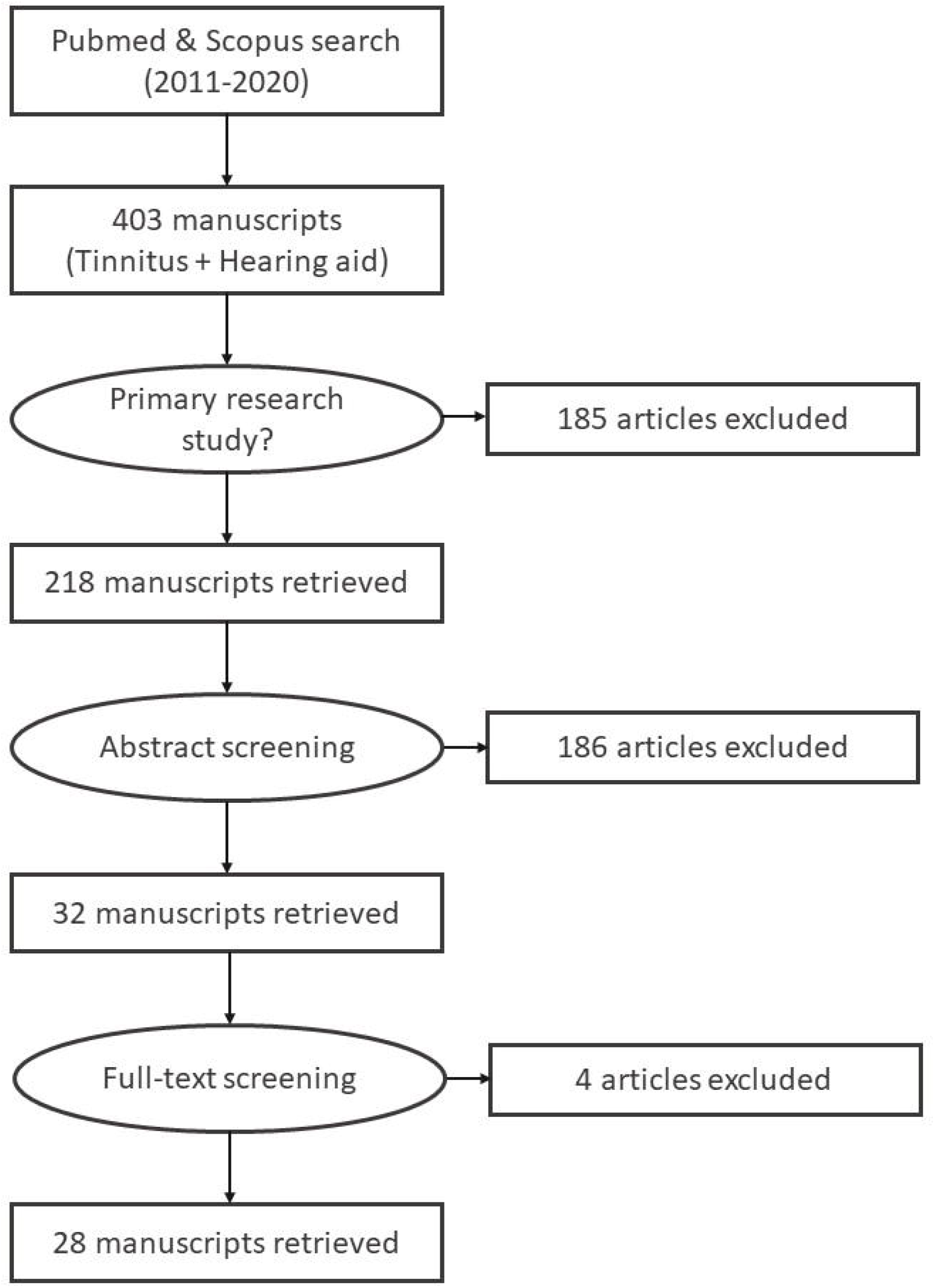
Chart of the percentage of studies presenting a significant, positive effect for tinnitus, no significant effect, or not applicable due to study aims.

## Results

The key elements of the 28 selected primary research studies are shown in chronological order in Table 1. The studies are extensively discussed in the current section. First, the studies focusing on the primary effect of sound amplification through a hearing aid are described, after which the studies investigating the additional effect of sound masking are presented.

**Table 1:** Overview of selected studies for this scoping review (HA, hearing aid; SG: sound generator)

| First author | Year | Control group | Fitting | SG? | Combined therapies | Sample size | Participants | Tinnitus effect? | Study design | Methodology |
| --- | --- | --- | --- | --- | --- | --- | --- | --- | --- | --- |
| Andersson | 2011 | No tinnitus | NA | No | No | 85 | HA users with and without tinnitus | NA | Retrospective | Cross-sectional, response rate 53%, low tinnitus distress |
| Peltier | 2012 | None | LOFT | No | No | 74 | Tinnitus, HF HL | No | Retrospective | No fixed time points, no clear inclusion and exclusion criteria |
| MCNeill | 2012 | None | NA | No | No | 70 | Bothersome chronic tinnitus | No | Retrospective |  |
| Oz | 2013 | Only betahistine | NAL-NL1, WB noise | Yes | Counseling | 21 | Primary complaint chronic tinnitus | Yes, no group differences | Prospective | RCT, all received betahistine, combination device or sound generator |
| Shekhawat | 2013 | None | DSL5 | No | No | 25 | HA candidates, chronic tinnitus | NA | Experimental | One time point, no follow up |
| Shekhawat | 2014 | Sham tDCS | DSL5 | No | tDCS | 40 | Chronic tinnitus | Yes | Prospective | Same HA, no washout period |
| dos Santos | 2014 | Conventional | NAL-NL1, White noise | Yes | Counseling | 49 | Chronic tinnitus and mild to moderate bilateral SNHL | Yes, no group differences | Prospective | Blind RCT, Same HA |
| Hodgson | 2015 | WDRC | DSL5, FC | No | Counseling | 16 | Chronic tinnitus and HF SNHL | Yes, no group differences | Prospective | Single-blind crossover trial |
| Henry | 2015 | Conventional | NAL-NL2, AM and FM noise | Yes | Counseling | 30 | Bothersome tinnitus, hearing candidate | Yes, no group differences | Prospective |  |
| Strauss | 2015 | Conventional | Notch | No | TRT | 20 | Tonal tinnitus | No | Prospective | Double-blind study, objective outcome measure |
| Jalilvand | 2015 | Conventional | NAL-NL1 | Yes | No | 974 | Blast-induced chronic tinnitus | NA | Longitudinal | All started with combination device |
| Araujo | 2016 | No tinnitus | NA | No | Tinnitus counseling | 24 | Elderly, moderate SNHL, with and without tinnitus | Yes | Prospective | Same HA, only women with tinnitus |
| Zarenoue | 2016 | No MI | NA | No | No | 46 | Tinnitus, SNHL | Yes, larger effect for MI* | Prospective | RCT, no stratification |
| Cabral | 2016 | None | NA | No | No | 17 | HA users, tinnitus | Yes | Observational | Not all persistent tinnitus (only 58%), different questionnaires pre vs. post |
| Sereda | 2016 | None | NA | Yes | No | 8 | Experienced, satisfied combination device users, chronic tinnitus | NA | Prospective | Same HA |
| Searchfield | 2016 | TRT-based sound therapy | DSL5, 3D masking | Yes | Counseling | 9 | Chronic unilateral tinnitus, mild to moderate HF SNHL | Yes, no group differences | Prospective | Same HA, cross over trial |
| Berberian | 2016 | None | NA | Yes | No | 25 | Bilateral HA users + SG with bilateral tinnitus, no severe to profound HL | Yes | Retrospective |  |
| Rocha | 2017 | Sound generator | NAL-NL1, White noise | Yes | Counseling | 30 | Bilateral chronic tinnitus | Yes, no group differences | Prospective | Same HA brand and model |
| Zarenoue | 2017 | No tinnitus | NA | No | No | 92 | Mild to moderate SNHL, with and without tinnitus | Yes | Prospective | Motivational interviewing was delivered to some patients (Zarenoue et al, 2016) |
| Tyler | 2017 | None | Zen tones | Yes | Zen Tinnitus Therapy | 20 | Chronic tinnitus | Yes | Prospective | Same brand, not everybody same trajectory, n = 7 already HA before study |
| Henry | 2017 | Conventional | NAL-NL2, EHWA | Yes | Counseling | 55 | Mild to moderately-severe hearing loss, bothersome tinnitus | Yes, no group differences | Prospective | RCT, same brand |
| Shabana | 2018 | Conventional | Zen tones | Yes | Counseling, Relaxation | 40 | SNHL, chronic bothersome tinnitus | Yes, larger effect for Zen tones* | Prospective | Monoaurally fitted, same brand |
| Yakunina | 2019 | WDRC | FT vs. LFT | No | No | 94 | Chronic tinnitus, SNHL | Yes, no group differences | Prospective | Double-blind RCT, same brand |
| Shetty | 2019 | None | NAL-NL2 or DSL5 | No | No | 20 | Catastrophic tinnitus (even with HA), bilateral SNHL | NA | Prospective | Test of tinnitus suppression during fitting, SPIN material validated? |
| Haab | 2019 | Conventional | Notch | No | No | 34 | Subjective chronic tonal tinnitus, mild to moderate HL | Group differences at 6m* | Prospective |  |
| Yokota | 2020 | None | NA | No | No | 66 | Primary complaint hearing disturbance, tinnitus | Yes | Retrospective |  |
| Marcum | 2020 | Conventional | Notch | No | No | 39 | Primary complaint of stable, tonal tinnitus, bilateral mild-to-moderate SNHL | No | Prospective | Double blind |
| Rocha | 2020 | None | NAL-NL1, White noise | Yes | Counseling | 40 | Chronic bilateral tinnitus and symmetrical bilateral mild to moderate SNHL | Yes | Prospective | Same HA |

### Hearing Aids

A total of 16 articles focused on fitting of hearing aids in a tinnitus population.

### Benefits of hearing aid use

The additional benefit for hearing aid users with tinnitus compared to hearing aid users without tinnitus was the topic of three studies included in this scoping review. Andersson et al. (2011) described retrospectively the benefits of hearing aid use in 52 tinnitus patients and 33 non-tinnitus patients. The tinnitus group reported more aversiveness and less benefit in difficult situations with a hearing aid. As the results were mainly from patients with a low tinnitus distress, extrapolation to the tinnitus population needs caution. In addition, the low response rate (53%) might have biased the results. Furthermore, a comparison between tinnitus (n=12) and non-tinnitus patients (n=12) after hearing aid use was made by Araujo et al. (2016) in an elderly population. The psychoacoustic measures of tinnitus, as well as the self-report questionnaires were assessed after 1 and 3 months of hearing aid use. The tinnitus loudness reduced significantly over time, as well as the nuisance with tinnitus and hearing loss. Moreover, all participants were satisfied with the use of hearing aids. However, tinnitus counselling was also provided during this study. Remarkably, all tinnitus patients were female. This potential gender bias might have influenced the results as it has been suggested that male and female patients respond differently to tinnitus treatments (Van der Wal et al. 2020). The most recent study on this topic was done by Zarenoe et al. (2017), who compared the effect of hearing aids on working memory, sleep and hearing problems. Their sample consisted of 46 tinnitus patients and 46 non-tinnitus patients who were followed-up until 3 months after completion of fitting in a prospective study. The tinnitus group showed a significantly larger improvement in terms of working memory (measured by the Reading Span) and self-reported sleep difficulties. Moreover, significant improvements in tinnitus distress were shown. While Araujo and Iório (2016) and Zarenoe et al. (2017) suggest a positive effect of hearing aid use on tinnitus, Andersson et al. (2011) call attention to the fact that hearing aid fitting might be more difficult in a tinnitus population due to aversiveness of sound. Moreover, the demonstrated positive effects on tinnitus can be mediated by the decrease in nuisance with hearing loss, improved sleep quality and improved cognitive function.

The secondary benefits in hearing-aid users who experience tinnitus were further investigated by Cabral et al. (2016) and Yokota et al. (2020). Cabral et al. (2016) showed significant changes in emotional and auditory complaints after three months of hearing aid use in 17 patients, however different questionnaires were applied before and after hearing aid use, which impedes interpretations of these results. The retrospective study by Yokota et al. (2020) evaluated changes in tinnitus distress before and 12 months after hearing-aid use in 66 patients with a primary complaint of hearing disturbance and co-existing tinnitus. Significant improvements were salient, even in patients with bilateral tinnitus who were fitted with a unilateral hearing aid. But for patients with unilateral tinnitus, there was no significant improvement in case the hearing aid was worn on the contralateral side. These two studies show that tinnitus relief can be apparent, even if that was not the primary motivation for the hearing aid use.

### Fitting adjustments for tinnitus relief

The effect of notch filter amplification was evaluated in three studies, all comparing this approach to conventional amplification. Two of the three studies did not report significant effects of either treatment on tinnitus perception. Strauss et al. (2015) included 20 patients with tonal tinnitus in a 3 weeks double-blind study. While more prominent improvement was shown for the notch-induced lateral inhibition, the authors did not report whether this result was significant. A more recent study by Marcrum et al. (2020) conducted a comparable study with 39 adults with a primary complaint of tonal tinnitus over a period of 12 weeks. The authors reported a minimal effect of either treatment on tinnitus symptoms. However, there was no significant difference between notch filter and conventional amplification (23.8% and 33.3% meaningful benefit according to Tinnitus Handicap Inventory; THI). Yet, the group of Strauss recently conducted a second study with notched hearing aids and were able to show significant improvements (Haab et al. 2019). The goal was to evaluate this treatment on the long-term (i.e. 6 months) with a control group being fitted with standard hearing aids. The group with notched hearing aids improved significantly in their Tinnitus Questionnaire (TQ) in comparison to the patients in the control group. In sum, notched acoustic stimulation was successfully integrated into patient’s daily routine and might be promising. However, further studies are required to replicate these recent results. It has to be noted that a precise and robust characterization of the patients’ tinnitus frequency and its’ possible fluctuations over time is crucial for therapeutic success.

As tinnitus is often associated with a descending-slope type of hearing loss (i.e. the most common type of hearing loss), signal processing strategies improving the audibility of high-frequency sounds have been evaluated in a tinnitus population. The retrospective study of Peltier et al. (2012) described the effect of Linear Frequency Transposition (LFT) in terms of tinnitus suppression for 74 patients. A total of 81% reported tinnitus suppression. However, it was not reported whether this suppression was significant. There were also no fixed time points for data collection, neither clear inclusion nor exclusion criteria. The effect of another frequency lowering technique, namely Frequency Compression (FC), was evaluated and compared to the conventional Wide Dynamic Range Compression (WDRC) in a crossover trial by Hodgson et al. (2015). While both treatments resulted in tinnitus improvements, WDRC showed larger improvements. It is important to note that every patient also received one counselling session. More recently, Frequency Translation (FT), LFT and WDRC were compared in a randomized double-blind controlled trial by Yakunina et al. (2019). A total of 94 tinnitus patients were randomized into the three groups and wore the hearing aid for three months, followed by a period of cessation for 3 months. The tinnitus perception improved for the groups post-intervention and at 3 months follow-up. However, no difference between the groups was found. Furthermore, the authors stated that the effect lasted for 3 months after stopping hearing aid use, but no analyses were conducted to compare the tinnitus perception at post intervention and follow up. While these three studies reported positive effects of frequency lowering techniques to some extent, no superiority over conventional amplification could be demonstrated.

Shekhawat et al. (2013) and Shetty and Pottackal (2019) focused on the fitting parameters preferred by patients for tinnitus relief. The study by Shekhawat et al. aimed to identify the optimized high frequency amplification for a first-fit. A total of 25 chronic tinnitus patients were asked to compare 13 speech files (i.e. simulating effects of a change in DSL i/o v5 prescription) in terms of their tinnitus perception. Overall, a reduction in output was most preferred. More specifically, A 6 dB reduction to the prescribed gain at 2 kHz was most preferred. There was a trend observed in which lower tinnitus pitch was associated with a preference for reduction in gain, but it failed to reach statistical significance. These findings may be limited as it was a lab experiment, and as such no follow up data on the effect of these prescriptions in daily life were available. More recently, the gain needed for tinnitus suppression was investigated in a more extensive study design which was applied by Shetty & Pottackal (2019). The effect on tinnitus perception and speech understanding in noise (SPIN) was evaluated up to 30 days of hearing aid use. Their sample consisted of a very specific group, namely 20 patients who experience catastrophic tinnitus (i.e. always heard, disturbed sleep pattern and difficulty with any activity) even after being fitted with a hearing aid. A comparison between the fitting formula DSL i/o v5 and NAL-NL2 showed that higher gain at the tinnitus pitch was needed for the latter formula in terms of tinnitus suppression. Interestingly, SPIN was not affected by tinnitus pitch or revised fitting formula. Moreover, these authors also found that a lower tinnitus pitch was associated with less gain needed, which is in agreement with the results by Shekhawat et al. (2013), and this association was significant. Also, in this study the gain adjustments were performed in the lab. Hence, authors of both studies recommended adjusting the gain at the tinnitus pitch using DSL (I/o) v5 for tinnitus management.

### Predictive factors for hearing aid use resulting in tinnitus relief

McNeill et al. (2012) evaluated retrospectively predictive factors for tinnitus relief in 70 patients with bothersome chronic tinnitus. The reduction in tinnitus complaints was significantly larger when participants achieved masking (i.e. hearing tinnitus softer or not at all), and masking was more likely in patients with good low-frequency hearing and a tinnitus pitch in the frequency range of the hearing aids. It is important to bear in mind that the ‘no masking’ group also had a lower tinnitus severity prior to the study. Clinical trials focusing on predictive factors for therapeutic success with hearing aids are warranted, as the tinnitus population is highly heterogeneous (Cederroth et al. 2019).

### Enhancement of sound therapy from hearing aids

Whether a combination of two existing treatments for tinnitus lead to a larger benefit was investigated by Shekhawat et al. (2014) by applying multisession transcranial direct current stimulation (tDCS) on five consecutive days before hearing-aid use.A total of 40 chronic tinnitus patients were enrolled in this 7-month during double-blind randomized clinical trial. The patients were randomized in two groups: sham vs. experimental tDCS. As the results showed a significant similar reduction in tinnitus severity for both groups, the hearing aid effects appeared independent of tDCS. Only one parameter changed after experimental tDCS, namely minimum masking levels. However, as hearing aids were fitted immediately following the tDCS sequence and the hearing aids had a strong effect, any tDCS effect might have been washed out by larger effects of sound amplification. It was recommended more recently to have a washout period of a few days between the stimulation sessions because the impact of multiple tDCS sessions on tinnitus being non-linear in nature (Shekhawat et al. 2018). Zarenoe et al. (2016) looked also into the optimization of hearing aids for tinnitus relief by adding motivational interviewing (MI) to their protocol. This pilot RCT included 50 patients with tinnitus and sensorineural hearing loss, but four patients dropped out due to dissatisfaction of hearing aid amplification. Hence, 23 patients received a brief MI program as an adjunct to hearing aid rehabilitation, whereas the other patients underwent conventional hearing aid fitting. The authors succeeded in their aim as a significant larger improvement was demonstrated for the first group 3 months after completing hearing aid fitting. While these results were positive, they might have been biased by differences in hearing thresholds and tinnitus distress at baseline between both groups. It was not clear if these differences were significant. These preliminary, but promising results, should be replicated in larger trials.

### Overview of results with amplification

In summary, out of the 16 studies described above, nine studies showed significant improvements in tinnitus perception (Araujo and Iório 2016; Cabral et al. 2016; Haab et al. 2019; Hodgson et al. 2015; Shekhawat et al. 2014; Yakunina et al. 2019; Yokota et al. 2020; Zarenoe et al. 2017; Zarenoe et al. 2016), while four studies did not report significant tinnitus relief (Marcrum et al. 2020; McNeill et al. 2012; Peltier et al. 2012; Strauss et al. 2015). The other three studies did not test whether hearing aids’ use leads to tinnitus relief (Andersson et al. 2011; Shekhawat, Searchfield, Kobayashi, et al. 2013; Shetty and Pottackal 2019).

### Additional masking in hearing aids

A total of 12 articles focused on adding an additional masking function to the hearing aids in a tinnitus population.

### Evaluation of combination devices

#### Additional benefit of sound generators in a hearing aid

A total of 3 studies focused on the effect of combination devices (i.e. amplification and sound generator) compared to conventional amplification. In 2014, dos Santos et al. investigated if combination devices (HA + SG) were more effective than conventional amplification alone (HA) (dos Santos et al. 2014). The study design was a blind randomized clinical trial that included 49 chronic tinnitus patients with a bilateral mild to moderate SNHL. The authors specified that the white noise in the sound generator was set at the lowest intensity capable of providing tinnitus relief and that the noise reduction strategies were switched off. Both groups showed significant improvements after 3 months of hearing aid use. Yet, there was no statistically significant difference between both groups. More specifically, 62.5 % of the first group (HA + SG) and 78 % of the second group (HA) showed a clinically significant reduction in tinnitus annoyance (i.e. a minimal reduction of 20 points on the THI). Notably, all participants used the same hearing aids, which were developed by the University of São Paulo. One year later, a study was published by Henry et al. (2015), in which a commercially available combination hearing aid (with an amplitude and frequency modulated noise stimulus) was evaluated in a study funded by Starkey Hearing Technologies. The 30 participants with bothersome tinnitus were hearing aid candidates who did not wear hearing aids in the previous 12 months. Similar to the previous study, both groups demonstrated significant improvements in terms of tinnitus distress, without a significant difference between groups. However, this study showed a trend that prioritized combination devices. Interestingly, the participants also filled out the self-report questionnaires for the situations in which they were not wearing the hearing aids. The results showed also significant improvements for those moments. Moreover, the sound generators did not result in any negative effect on hearing handicap. Similar results were found by Henry et al. (2017), who replicated this study, although the devices used were from a different manufacturer (i.e. Phonak, with three SG options: white noise, pink noise and spectrally shaped noise) and a third comparison option was added (i.e. extended-wear, deep fit hearing aids; EWHA – worn 24h/day, 7d/week). The inclusion of EWHAs was based on observations that many patients using this type of hearing aid perceived their tinnitus as less bothersome. This RCT included 55 patients with a mild to moderately-severe hearing loss and bothersome tinnitus. Moreover, hearing-specific questionnaires and a speech-in-noise test showed similar improvements in the three groups. Although not significant, the tinnitus relief was smaller for the HA only group and the hearing improvement was smaller for the EWHA group. In sum, a superiority of combination devices compared to conventional hearing aids has not been proven. On the other hand, there is also no inferiority of combination devices in terms of speech understanding demonstrated in these papers.

The effectiveness of a combination device (HA + SG) for 15 patients with a hearing loss was compared to a sound generator only (SG) for 15 normal hearing patients (NH) in a study by Rocha et al. (2017). All patients had bilateral chronic tinnitus, used the same hearing aid with a constant white noise and participated in a counselling session. This approach was successful as both groups revealed significant improvements at six months follow-up. The authors concluded that the use of a sound generator was similarly effective for NH patients as a combination device for patients with a hearing loss. However, the results might be yet again mediated by the effect of counselling.

The satisfactory rates of SG, HA or SG + HA were investigated in a longitudinal study specifically focusing on war veterans with blast-induced chronic tinnitus (Jalilvand et al. 2015). More specifically, 974 patients were provided with a sound generator and a hearing aid at baseline and were followed up for two years. The satisfaction scores for HA and HA + SG increased with time, while the scores decreased for SG. This could be explained by patients tolerating the amplification better over time, resulting in more amplification and higher satisfaction rates. On the other hand, the sound generator gradually became unpleasant for the patients, according to the authors. Most patients reported to prefer a hearing aid only (HA: 84 %, HA + SG: 13.3 %, SG: 2.7 %). It is important to note that in the early years of this study (2004), hearing aids and sound generators were fitted as separate devices. As such, the number of subjects who preferred both in those times was very small. The authors did not find explaining variables such as audiological parameters or tinnitus characteristics.

While most studies include a control group who receive conventional amplification, sound generator, counselling or no therapy at all, Oz et al. (2013) administered betahistine to 21 patients with a primary complaint of chronic tinnitus in an RCT. A total of 12 patients received in addition a combination device or a sound generator (both with a wideband noise), depending on the hearing loss. Both groups showed positive effects on tinnitus after 3 months. It has to be noted that this study claims to be double blinded, but it is unclear how they blinded patients for using a hearing aid. Moreover, a recent Cochrane review shows that there is no evidence to suggest that betahistine has an effect on subjective idiopathic tinnitus (Wegner et al. 2018).

#### Benefit in terms of hearing and tinnitus

Two studies included in this scoping review did not include a control group in the evaluation of combination devices. Berberian et al. (2017) evaluated bilateral use of combination devices for 6 months in 25 patients with bilateral tinnitus and mild to moderately severe hearing loss. They showed significant improvements in hearing thresholds and tinnitus perception. Similarly, Rocha & Mondelli (2020) evaluated the benefit of combination devices (with white noise) over 6 months. A total of 40 chronic bilateral tinnitus patients with symmetrical bilateral mild to moderate SNHL were included. Real ear measurements (REM) were performed to verify the sound generator. Significant tinnitus improvements were shown. Hence, these two studies are in agreement with previously described studies as they show positive effects of combination devices but could not contribute to the discussion whether additional masking is beneficiary.

### Sound generator, masking, .. What’s in a name?

While many studies focus on the effect of sound generators and combination devices, some studies focused more on the ideal sound stimulus to provide tinnitus relief. Searchfield et al. (2016) investigated thoroughly masking at the perceived spatial location of the patient’s tinnitus (i.e. 3D masking). Their paper describes three studies: a proof-of-concept study, a prototype evaluation, and a four-month crossover pilot study. A preference for the 3D masking stimulus and less preference for unilateral masking was shown. Moreover, the 3D masking did not have to be as loud as other maskers. Finally, the prototype evaluation demonstrated significant larger tinnitus improvements with the 3D masking.

A sound therapy introduced by Widex is called Zen Tones, which uses music that is generated based on a fractal algorithm. Adjustments can be made in terms of volume, pitch, and tempo. Moreover, automatic adjustments are made based on the ambient noise in the environment. A pilot study with the Widex Zen Tinnitus Therapy was performed in 2017 by Tyler at al., evaluating the progression of benefits in 20 chronic tinnitus patients receiving counselling, hearing aids, tinnitus activities and Zen therapy. They demonstrated that all participants significantly benefited from this approach. Large benefits were observed following the last two elements of the treatment. It is important to bear in mind that seven patients were already hearing aid users, and that not all patients needed the same amount and type of appointments. An evaluation of the additional benefit of Zen tones was conducted by Shabana et al. (2018) with 40 chronic tinnitus patients with a SNHL. Also, in this study the patients followed first 2 months of counselling before using hearing aids for 4 months. Zen tones were made available for 20 participants (i.e. study group). Significant improvements were only demonstrated after counselling and amplification. Yet, a significantly greater benefit was apparent for the study group. It has to be noted that all patients were fitted monaurally for financial reasons.

A pre-market version of Oticon Alta with a tinnitus sound generator was evaluated in a feasibility study of Sereda et al. (2017). The eight participants, who were experienced and satisfied hearing aid users with chronic tinnitus, could select different types of noise (i.e. white/pink/brown, unmodulated or modulated, non-filtered or bandpassed) and three different ocean sounds. Participants reported to be satisfied with the feeling of control and the variations possible. As can be expected, preferences depended on the individual and the situation. Yet, participants reported that the broadband noise was the most effective masker and the ocean noises were more distracting and/or relaxing. In general, combination devices were equally preferred over the conventional amplification.

### Overview of results with additional masking

Tinnitus complaints significantly decreased in ten studies (Berberian et al. 2017; dos Santos et al. 2014; Henry et al. 2015; Henry et al. 2017; Oz et al. 2013; Rocha and Mondelli 2017, 2020; Searchfield et al. 2016; Shabana et al. 2018; Tyler et al. 2017). Two studies did not conduct analyses to investigate if hearing aid use and/or the masking function result into tinnitus relief (Jalilvand et al. 2015; Sereda et al. 2017).

## Discussion

In view of all that has been mentioned so far, one may suppose that hearing aid use improves tinnitus perception. However, the quality of evidence could be questioned. Nevertheless, recent studies were more likely to focus on (1) optimizing the effect of hearing aids, by investigating different forms of amplification, and (2) better predicting which tinnitus patients benefit from hearing aids.

When optimizing tinnitus relief through hearing aid use, it is important to bear in mind that hearing aid fitting for chronic tinnitus patients can be challenging, as they require different fitting strategies. These patients might show increased aversiveness and decreased benefit in difficult listening situations with a hearing aid (Andersson et al. 2011). Decreased gain might be needed, specifically with patients who experience a lower tinnitus pitch. More advanced strategies, such as notch filter amplification and frequency lowering, demonstrate promising results, though the added benefits are still debated. Similarly, the addition of sound generators in a combination device has resulted in tinnitus improvement without a clear added benefit compared to conventional amplification. Jalilvand et al. in 2015 stated that amplification might be better tolerated over time, while sound generators might gradually become unpleasant (Jalilvand et al. 2015). The ideal noise settings probably depend on the individual patient and the situation. Furthermore, while there is a general preference for the use of binaural hearing aids in tinnitus patients, monaural hearing aids have also provided positive results, calling attention to cost-efficiency (Shabana et al. 2018; Yokota et al. 2020).

Predicting those who can most benefit from hearing aids in terms of tinnitus experience remains a challenge. Mc Neill et al. in 2012 found that patients with good low-frequency hearing and a tinnitus pitch in the frequency range of the hearing aids might perceive a larger tinnitus improvement (McNeill et al. 2012). It is clear that, as most studies apply different amplification protocols and reach significant improvements, more research is needed to uncover what works for whom.

The use of digital noise reduction and adaptive directional microphone systems has also been discussed in some studies, as they might reduce the potential impact of hearing aid use on tinnitus severity. The idea behind switching these digital features off is that continuous exposure to ambient noise levels may lead to reduction of gain in the auditory pathway. However, these features might be needed for beneficial effects in terms of hearing (dos Santos et al. 2014; Marcrum et al. 2020; McNeill et al. 2012; Shekhawat, Searchfield, Kobayashi, et al. 2013; Shetty and Pottackal 2019). Unfortunately, this topic was not further investigated since 2011 to our knowledge.

Another ongoing debate concerns the degree of tinnitus complaints when the hearing aid is switched off (for a few hours or even longer). While this topic was also not specifically investigated in the retrieved studies, some authors have discussed the matter. Most of them reported smaller positive results without the hearing aid device (Araujo and Iório 2016; Henry et al. 2015). Nevertheless, Yakunina et al. (2019) reported that the tinnitus improvements lasted for at least 3 months after 3 months of hearing aid use. Hence, these authors suggest that the mechanism of tinnitus suppression is beyond temporary masking and distraction. Another possible explanation is the effect of counseling, which was often part of the treatment in the reported studies, and this effect is not dependent on wearing the hearing aid or not. While for research purposes including counseling influences the results, in clinical practice it is often part of the multidisciplinary approach.

The findings of the current scoping review may be somewhat limited by the quality of the presented studies. First, some studies did not investigate the standalone effect of hearing aids, as they provided other treatments during the study (e.g. counselling) and/or did not include a control group. Moreover, some studies analyzed clinical gathered data, without a prospective fixed protocol, which impedes the formation of strong conclusions.

### Consultation exercise

Practitioners and patients’ view on the current topic are a final, but valuable, stage of the current scoping review. In a Delphi review of 2015, including 19 UK hearing professionals, Sereda and colleagues found that the presence of bothersome tinnitus supported the choice for hearing aids in patients with hearing difficulties and realistic expectations of this technology. Moreover, ‘open-fit technology’ and ‘bilateral fitting’ were part of the usual care for bothersome tinnitus. However, the authors stated that a clear link between diagnostic information and recommended treatment was lacking, as well as specific guidelines and recommended questionnaires (Sereda et al. 2015). For the current study, seven experts in the field (i.e. 3 audiology researchers, 1 clinical audiologist with expertise in hearing aid fitting for tinnitus, 1 tinnitus consultant and 2 representatives of a patient association) were consulted about the study findings. In general, the findings were in agreement with their view on the scientific evidence of hearing aids for tinnitus and they also emphasized the importance for RCTs in order to reach consensus. The input of the experts is summarized in the following points:

1. The high variability in effects on tinnitus perception is similar to the experiences in clinical practice, especially at first. Fitting and adjusting seems to take longer in this population. It is important to further explore which patients can benefit from hearing aid fitting (e.g. role of age, tinnitus pitch, personality, distress, sleeping problems, coping, motivation, expectations) and what ‘motivational interviewing’ can contribute to this issue. Moreover, several questions remain on the fitting method itself (e.g. importance of comfort, sound quality, REM). Consequently, this could support development of guidelines for clinicians, as well as for patient associations.
2. The use of sound generators is often temporarily and in combination with psychoeducation. Patients often report the relaxation effect of these additional sounds. Hence, the question arises which sound stimuli should be used (e.g. music, nature sounds, pure stimuli, etc). With regards to combination devices, datalogging is needed to uncover how long patients use different programs and in which situations. As patients often report an aggravation of tinnitus after switching the masking off, the risk of sound generators interfering with the habituation process came up in this consultation phase and should be the topic of future research.
3. As counseling is often an essential part of a multidisciplinary approach, the experts recommended future studies to focus on the content and the extent of the counseling needed in a hearing aid fitting. Excluding counseling might lead to counterproductive effects in their opinion.
4. More attention should be given to the different hearing losses (e.g. type and degree of hearing loss) and the effects of different hearing solutions needed, as well as the relationship between the change in audibility/listening effort and the change in tinnitus perception.

## Conclusions

As there is a long history of clinical work and research on hearing aids for tinnitus relief, there was a need for recent literature to be reviewed and guide decision making in tinnitus management. Moreover, a consultation exercise, which is an essential stage of a scoping review, was included. The majority of the studies supported the use of hearing aids for tinnitus relief, however, the quality of evidence is low, limiting the confidence in the scientific support. The experts stated that the findings were in agreement with their view on the scientific evidence and they emphasized the importance of reaching consensus. While a stronger methodology is recommended, future studies should determine the optimal hearing solution for sound amplification and sound generation in tinnitus patients. More attention needs to be brought on how it should be fitted and the ideal parameters, as well as on how this can be part of a larger multidisciplinary approach. Finally, the underlying working mechanisms must be further explored, more specifically in order to understand the effect of switching off the hearing aid at night and the predictive factors for therapy success.

## Data Availability

All data is shown in the manuscript itself, as it is a scoping review.

## ACKNOWLEDGMENTS

We gratefully acknowledge the valuable input of the experts of the consultation stage, Prof. dr. David Baguley, drs. Rene van der Wilk, Nancy Van Looveren, Prof. dr. Vinaya Manchaiah, Prof. dr. Annick Gilles, Bert Lecomte and Wout De Coster.

